# Genome-wide association study and multi-trait analysis of opioid use disorder identifies novel associations in 639,709 individuals of European and African ancestry

**DOI:** 10.1101/2021.12.04.21267094

**Authors:** Joseph D. Deak, Hang Zhou, Marco Galimberti, Daniel Levey, Frank R. Wendt, Sandra Sanchez-Roige, Alexander Hatoum, Emma C. Johnson, Yaira Z. Nunez, Ditte Demontis, Anders D. Børglum, Veera M. Rajagopal, Mariela V. Jennings, Rachel L. Kember, Amy C. Justice, Howard J. Edenberg, Arpana Agrawal, Renato Polimanti, Henry R. Kranzler, Joel Gelernter

## Abstract

**Background:** Despite the large toll of opioid use disorder (OUD), genome-wide association studies (GWAS) of OUD to date have yielded few susceptibility loci.

**Methods:** We performed a large-scale GWAS of OUD in individuals of European (EUR) and African (AFR) ancestry, optimizing genetic informativeness by performing MTAG (Multi-trait analysis of GWAS) with genetically correlated substance use disorders (SUDs). Meta-analysis included seven cohorts: the Million Veteran Program (MVP), Psychiatric Genomics Consortium (PGC), iPSYCH, FinnGen, Partners Biobank, BioVU, and Yale-Penn 3, resulting in a total N=639,709 (N_cases_=20,858) across ancestries. OUD cases were defined as having lifetime OUD diagnosis, and controls as anyone not known to meet OUD criteria. We estimated SNP-heritability (h^2^_SNP_) and genetic correlations (r_g_). Based on genetic correlation, we performed MTAG on OUD, alcohol use disorder (AUD), and cannabis use disorder (CanUD).

**Results:** The EUR meta-analysis identified three genome-wide significant (GWS; *p*≤5×10^−8^) lead SNPs—one at *FURIN* (rs11372849; *p*=9.54×10^−10^) and two *OPRM1* variants (rs1799971, *p*=4.92×10^−09^ ; rs79704991, *p*=1.37×10^−08^; r^2^=0.02). Rs1799971 (*p*=4.91×10^−08^) and another *OPRM1* variant (rs9478500; *p*=1.95×10^−8^; r^2^=0.03) were identified in the cross-ancestry meta-analysis. Estimated h^2^ _SNP_ was 12.75%, with strong r_g_ with CanUD (r_g_ =0.82; *p*=1.14×10^−47^) and AUD (r_g_=0.77; *p*=6.36×10^−78^). The OUD-MTAG resulted in 18 GWS loci, some of which map to genes or gene regions that have previously been associated with psychiatric or addiction phenotypes.

**Conclusions:** We identified multiple OUD variant associations at *OPRM1*, single variant associations with *FURIN*, and 18 GWS associations in the OUD-MTAG. OUD is likely influenced by both OUD-specific loci and loci shared across SUDs.

## Introduction

Opioid use disorder (OUD) has a serious negative impact on public health and is a leading cause of preventable death(1). Although opioid misuse and progression to OUD(2) are influenced by heritable factors, discovery of OUD risk loci has been limited(3-7). Difficulties in advancing OUD genetic discovery are largely due to lack of adequately powered cohorts of genetically informative samples(8,9).

Genome-wide association studies (GWAS) of OUD have been underpowered(8,9). Nevertheless, recent progress in GWAS of OUD include the identification and confirmation of a genome-wide significant (GWS) functional variant (rs1799971) in *OPRM1*(7). Earlier OUD GWAS identified associations with variation in several genes including *KCNG2, KCNC1, APBB2, CNIH3, RGMA*, and *OPRM1*(3-6), but the validity of those associations remains largely untested due to the lack of statistically powerful independent OUD cohorts. In addition to specific genetic loci, OUD GWAS have also demonstrated genetic correlations (r_g_) with other substance use disorders (SUDs) (e.g. alcohol use disorder [AUD]; r_g_=0.73) and psychiatric disorders (e.g. attention-deficit hyperactivity disorder; [r_g_=0.36])(7).

Large-scale GWAS meta-analytic techniques have proven valuable in advancing discovery of novel loci for other SUDs (e.g., AUD, problematic alcohol use (PAU), cannabis use disorder (CanUD)(10-12). This study applies similar meta-analytic methods for OUD by combining GWAS effects across multiple studies and two ancestral groups.

Multi-trait methods (e.g., MTAG; Multi-trait analysis of GWAS)(13) have the potential to increase power for OUD gene discovery. MTAG capitalizes on the r_g_ between genetically-related traits (e.g., r_g_ >0.70) to increase the equivalent sample size. MTAG is an attractive option for boosting power for sets of similar traits (e.g., SUDs)(11,14), and holds particular promise for disorders such as OUD for which only a limited number of cases are available for analysis. MTAG can generate estimates of trait-specific effects that leverage information from multiple GWAS summary statistics while accounting for both known and unknown sample overlap across the discovery samples(13). Thus, MTAG can maximize the genetic information available for OUD by leveraging the statistical power of GWAS of non-opioid SUDs, to advance our understanding of the genetic etiology of OUD and shared genetic liability across SUDs.

We conducted a large-scale GWAS meta-analysis of OUD in samples of African (AFR) and European (EUR) ancestry individuals. We maximized the informativeness of the available samples by performing a multi-trait analysis that incorporates SUDs that are highly genetically correlated with OUD.

## Methods

### Data and Participants

The GWAS meta-analysis includes summary statistics across seven cohorts examining OUD case vs. OUD control status in AFR and EUR ancestry individuals. We included both published and unpublished OUD GWAS. Previously published GWAS include data from Yale-Penn(3,6,7), PGC-SUD(6), and the Partners Biobank(15). For MVP Releases 1 and 2 (the data releases used in the present analysis), a previous GWAS of OUD cases vs. opioid-exposed controls was reported(7). MVP data included in the current meta-analysis use a different control definition (unscreened controls) to align better with the control definitions available in most other samples included in the meta-analysis. GWAS summary data for FinnGen(16) was accessed via a publicly available repository (https://r5.finngen.fi/). GWAS of OUD from iPSYCH(17), BioVU(18), and newly-available data from Yale-Penn subjects (Yale-Penn 3), previously unpublished, were performed by analysts at their respective study sites (**Supplemental Note**). We thus had a total AFR sample of 84,877 (N_case_=5,435 N_effective_=20,032), a total EUR sample size of 554,832 (N_case_=15,423; N_effective_=56,991), and an overall sample of 639,709 (N_case_=20,858; N_effective_=77,023). Other than Yale-Penn, this study involved de-identified data. The work was approved as appropriate by the Central VA institutional review board (IRB) and site-specific IRBs, including Yale University School of Medicine and VA CT, and was conducted in accordance with all relevant ethical regulations. Cohort-specific summaries of OUD cases and controls across AFR and EUR ancestry individuals are presented in **Table 1**. Additional phenotyping considerations are described in **Supplemental Note**.

**Table 1.** Overview of samples included in GWAS meta-analysis of OUD cases *vs*. OUD controls

| **Cohorts** | **EUR cases** | **EUR controls** | **AFR cases** | **AFR Controls** | **Case definition** | **Control definition** |
| --- | --- | --- | --- | --- | --- | --- |
| MVP Releases 1-2 | 8,529 | 267,737 | 4,032 | 71,511 | ICD-9/ICD-10 | unscreened |
| PGC-SUD* | 3,444 | 25,911 | 1,231 | 7,063 | DSM-IV | unexposed |
| Partners Biobank | 1,039 | 20,271 | - | - | ICD-9/ICD-10 | no SUD diagnosis |
| BioVU | 933 | 3,732 | - | - | ICD-9/ICD-10 | unscreened |
| FinnGen | 651 | 214,999 | - | - | ICD-9/ICD-10 | unscreened |
| Yale-Penn 3 | 448 | 1538 | 172 | 868 | DSM-IV | no OUD diagnosis |
| iPSYCH | 379 | 5221 | - | - | ICD-9/ICD-10 | no OUD diagnosis |
| **Ancestry-specific subtotals** | 15,423 | 539,409 | 5,435 | 79,442 |  |  |
| **Overall total cases** | 20,858 |  |  |  |  |  |
| **Overall total controls** | 618,851 |  |  |  |  |  |
| **Overall total N** | 639,709 | **Total N<sub>effective</sub>** | 77,023 |  |  |  |
| **EUR total N** | 554,832 | **EUR N<sub>effective</sub>** | 56,991 |  |  |  |
| **AFR total N** | 84,877 | **AFR N<sub>effective</sub>** | 20,032 |  |  |  |

\*The PGC-SUD OUD analysis included AFR and EUR participants from Yale-Penn 1 (N=3,922; Ncases=1,656) and Yale-Penn 2 (N=2,483; Ncases=846)—data from Yale-Penn supplied 67.4% of AFR cases and 51.1% of EUR cases for the overall PGC-SUD meta-analysis. Yale-Penn 3 is included as a separate cohort in the current analysis.

**Table 2.**
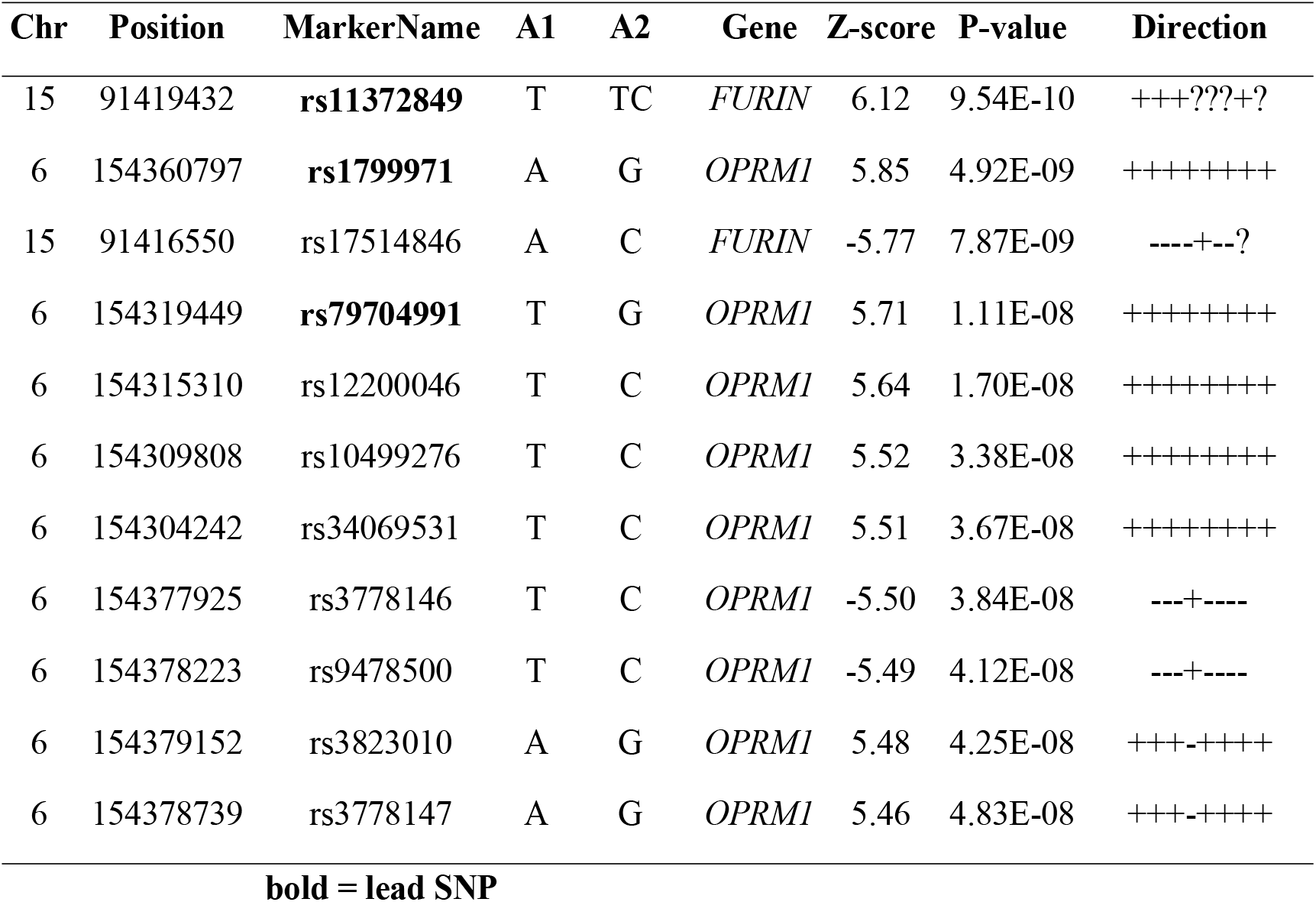
Genome-wide significant (*p*=5.00×10^−08^) GWAS associations in EUR OUD analysis.

**Table 3.**
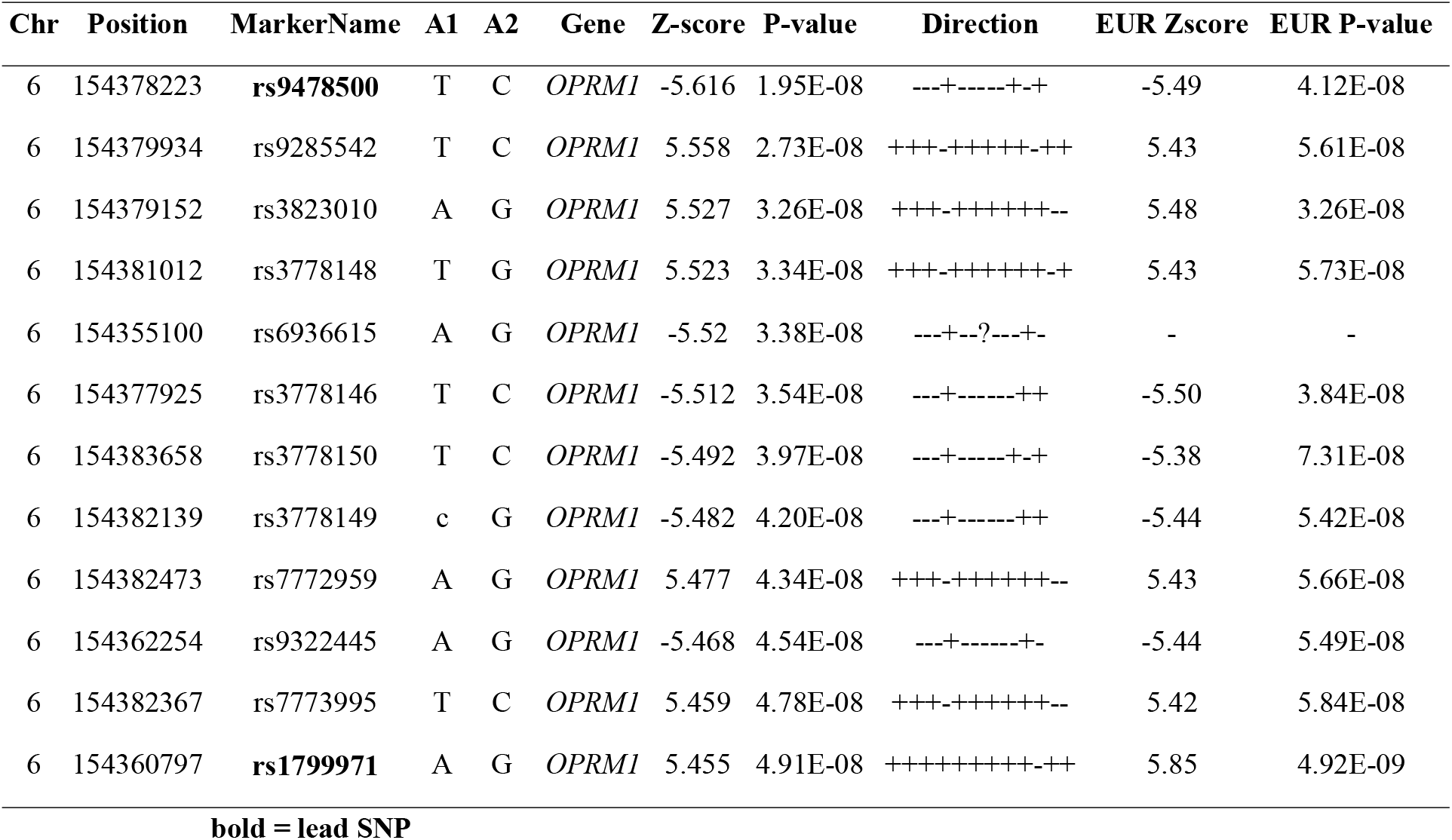
Genome-wide significant (*p*=5.00×10^−08^) GWAS associations in cross-ancestry OUD analysis.

### Ancestry-specific and cross-ancestry GWAS meta-analysis

GWAS samples were combined using an effective sample-size weighted meta-analysis in METAL (19). Ancestry-specific and cross-ancestry meta-analyses were performed. Measures of cross-sample heterogeneity (Cochran’s Q, I^2^) and genomic inflation (λ_GC_) were used to evaluate whether results were unduly influenced by heterogeneity between cohorts or by population stratification. GWAS summary statistics included in the meta-analysis were limited to variants present in at least 80% of the analysis-specific effective sample size (e.g., 80% of EUR N_effective_=45,593). The 80% effective sample size inclusion threshold ensured that variant effects present only in smaller cohorts did not disproportionately influence the overall results. This effectively meant that a variant needed to be present in MVP, PGC-SUD, and at least one additional cohort for it to be included in the meta-analysis. (**Figure 1**).

**Figure 1.**
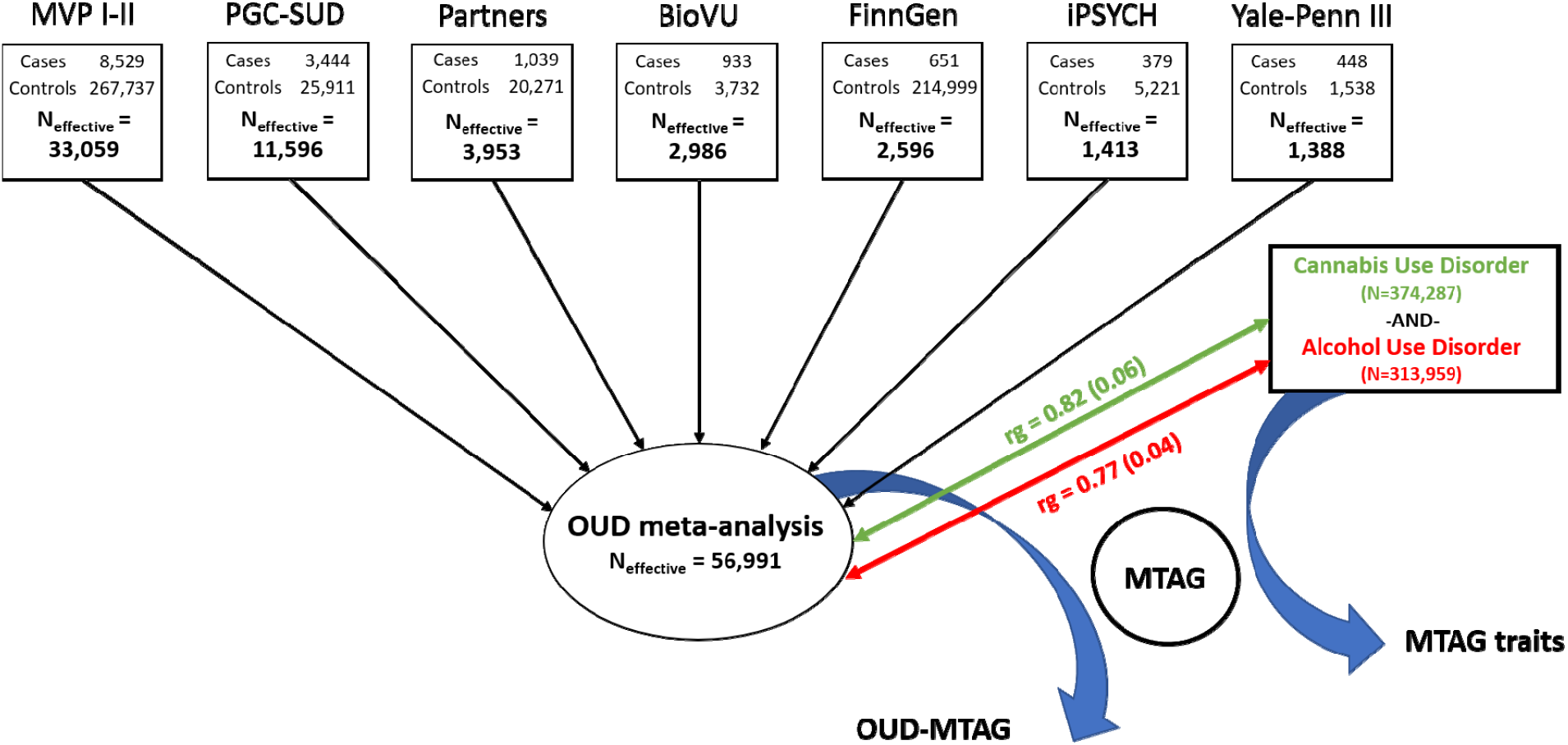
Overview of European-ancestry opioid use disorder (OUD) genome-wide association study and OUD multi-trait analysis.

Data from the 1000 Genome Project (1000G) phase 3(20) was used as a reference panel to determine EUR and AFR linkage-disequilibrium (LD) structure. Variants were mapped to the nearest gene based upon physical position (<10 kb from assigned gene). Conditional analyses were conducted using GCTA-COJO(21) to examine the conditional independence of genome-wide significant (GWS; *p*=5.00×10^−08^) of *OPRM1* variants in low LD (r^2^<0.1).

### SNP-heritability and Linkage-Disequilibrium (LD) Score Regression

GWAS summary statistics from the EUR OUD GWAS were used to estimate SNP-heritability (h^2^_SNP_) and to characterize OUD genetic correlations (r_g_) using LD score regression (LDSC)(22). LDSC analyses were restricted to HapMap3 variants(23). Effective sample-size was used in all LDSC-based analytic steps. Genetic correlations were estimated between OUD and other SUDs, traits related to substance use, psychiatric traits, chronic pain outcomes, sociodemographic factors, and additional traits of interest (**Supplemental Tables**). LDSC analyses were not performed in AFR and cross-ancestry meta-analyses because of the inability to use an LD reference panel for recently-admixed populations (e.g, African-Americans) and for analyses integrating datasets from diverse ancestry groups(22).

### Multi-trait analysis of GWAS summary statistics (MTAG)

Based on LDSC estimates of genetic correlations with OUD, a joint-analysis that included the EUR OUD GWAS and GWAS summary statistics for AUD(11) and CanUD(12) was conducted using MTAG(13). The AUD GWAS summary statistics used in the present analysis were made available as part of a broader GWAS of problematic alcohol use(11). MTAG was performed using study-specific effective sample sizes for the respective GWAS. Study-specific effect sizes were transformed to Z-scores so as to be on a uniform scale across the three GWAS included in the MTAG analysis. Genetic variants included in the MTAG analysis were filtered using default MTAG parameters(13). Briefly, variants were restricted to those common to all three of the GWAS, with a minor allele frequency (MAF) >0.01, and present in at least 75% of the 90th percentile of the study-specific SNP sample sizes. These MTAG parameters guard against heterogeneity in the distribution of common vs. rare variant effects, ensuring that SNP effects generated from relatively small subsets of the contributing discovery GWAS do not bias the effect estimates across traits(13).

### Phenome-wide Association Study (PheWAS)

To examine phenome-wide relationships for the EUR OUD GWAS and the OUD-MTAG analysis, and to compare their relationships with other clinically-relevant outcomes, we performed phenome-wide association studies (PheWAS) in BioVU(18). Briefly, BioVU is a cohort of >66,000 genotyped patients, with phenotypic data currently available for 1338 clinical outcomes from EHRs(18). Polygenic scores (PGS) for the EUR OUD GWAS and the OUD-MTAG analysis were computed using PRS-CS(24), excluding the subset of BioVU participants included in the EUR OUD GWAS. The respective PGS were then included in individual logistic regression models regressed on 1,291 clinical outcomes with case counts ≥100, covarying for sex, age, and the first 10 genetic PCs. Statistical significance for the PheWAS was defined as *p*=3.87×10^−05^ (0.05/1,291).

## Results

### Ancestry-specific and cross-ancestry GWAS meta-analyses

In the ancestry-specific analyses, there were three GWS risk variants (**Figure 2)** in the EUR GWAS. The top association was with a locus (rs11372849; *p*=9.54×10^−10^) located at the *FURIN* gene on chromosome 15, one of two GWS SNPs in that gene (rs17514846; r^2^=0.91). The second strongest association was with the *ORPM1* functional variant (rs1799971; *p*=4.92×10^−09^). An additional *OPRM1* variant was also identified (rs79704991; *p*=1.11×10^−08^; r^2^=0.02)(*OPRM1* regional plots—**Supplemental Figure S1**). GCTA-COJO (21) was used to conduct a conditional analysis of the two GWS *OPRM1* variants demonstrating low LD (rs1799971 conditioned on rs79704991 and vice versa). In these analyses, each variant fell below GWS when conditioning on the effect of the other (conditioned rs1799971-*p*_*conditioned*_=1.66×10^−06^; rs79704991-*p*_*conditioned*_=3.71×10^−06^); although, there were no statistically significant differences in effect estimates for the respective *OPRM1* variants conditioned vs. unconditioned effects. No GWS variants were identified in the AFR GWAS (**Supplemental Figure S2)**.

**Figure 2.**
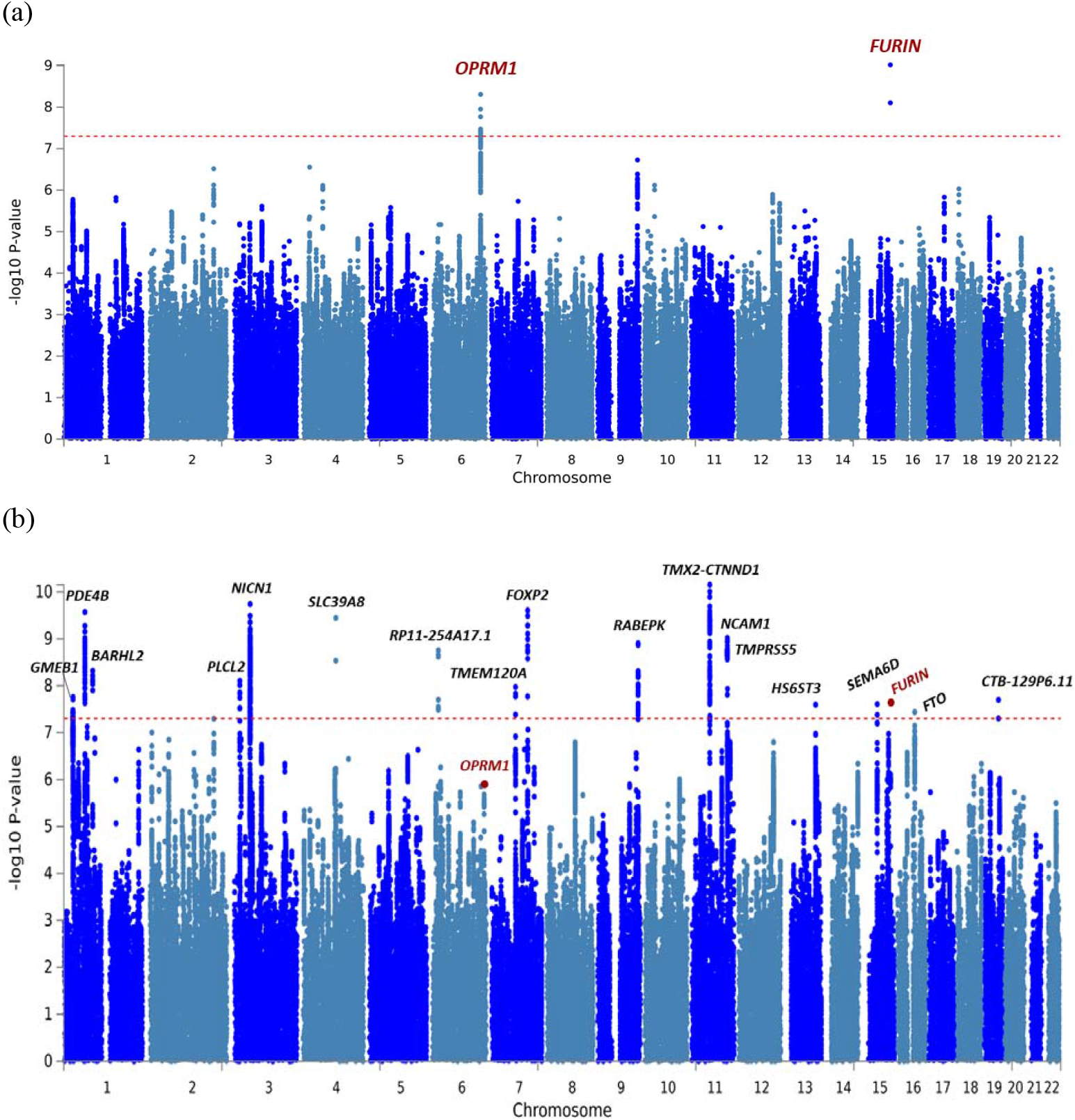
Manhattan plots of (a) European-ancestry OUD GWAS results and (b) OUD-MTAG multi-trait GWAS results.

The cross-ancestry OUD GWAS identified two GWS risk variants mapping to *OPRM1* (**Supplemental Figure S3**). The top association was with rs9478500 (*p*=1.95×10^−08^), an intronic variant. Rs1799971 was also GWS in the cross-ancestry meta-analysis (*p*=4.91×10^−08^), and is not in strong LD with rs9478500 (EUR r^2^=0.03; AFR r^2^=0.002; ALL r^2^=0.04). The top *FURIN* association in EUR (rs11372849) was uninformative in three of four AFR ancestry cohorts and did not meet the threshold (80% of N_effective_) we set to be included in the analysis. The second top *FURIN* association (rs17514846) fell just below GWS in the cross-ancestry GWAS (*p*=6.00×10^−08^).

### Gene and gene-set analysis

Gene and gene-set analyses are described in **Supplemental Note**. Both *FURIN* (*p*=3.09×10^−07^) and *OPRM1* (*p*=3.59×10^−07^) were significant in EUR gene-based analyses (**Supplemental Figure S4**). The EUR gene-set analysis resulted in a statistically significant set of 5 genes (*ANXA2, APOE, FURIN, MYLIP*, and *PCSK9*) involved in the regulation of the low-density lipoprotein (LDL) particle receptor catabolic process (*p*=2.79×10^−06^)(**Supplemental Tables**). No genes or gene-sets were GWS in the smaller AFR-specific analysis. In the cross-ancestry gene-based analysis, *FURIN* (*p*=6.00×10^−08^) and *OPRM1* (*p*=1.12×10^−07^) were significant (**Supplemental Figure S5**).

### SNP-heritability and Linkage-Disequilibrium (LD) Score Regression

For the EUR OUD GWAS, the liability scale SNP-heritability (h^2^_SNP_) estimate was 12.75% (s.e.=0.011) using effective sample-size adjusted prevalence rates and a population prevalence of 0.021(25). Genome-wide inflation was mild with respect to sample size and favored OUD polygenicity as indicated by the LDSC inflation factor (λ_GC_=1.18), intercept=1.01 (s.e.=0.011), and attenuation ratio=0.05 (s.e.=0.049).

In the EUR OUD GWAS, OUD showed statistically significant genetic correlations with substance use and SUDs, psychiatric traits, pain outcomes, physical health, and sociodemographic characteristics (**Figure 3**). The OUD trait in the current study was strongly genetically correlated with the largest published GWAS of OUD to date (r_g_=1.02; *p*=2.38×10^−214^)(7), suggesting that OUD is being captured consistently across the studies, as might be expected given the substantial overlap in OUD cases between the two studies, although the control definitions differed between analyses. OUD was also strongly genetically correlated with other SUDs, including CanUD (r_g_=0.82; *p* =1.14×10^−47^)(12) and AUD (r_g_=0.77; *p* = 6.36×10^−78^)(11). Modest genetic correlations were found for measures of substance use (e.g., the quantity/frequency alcohol use measure AUDIT-C) (r_g_=0.14; *p*=8.15×10^−03^)(10).

**Figure 3.**
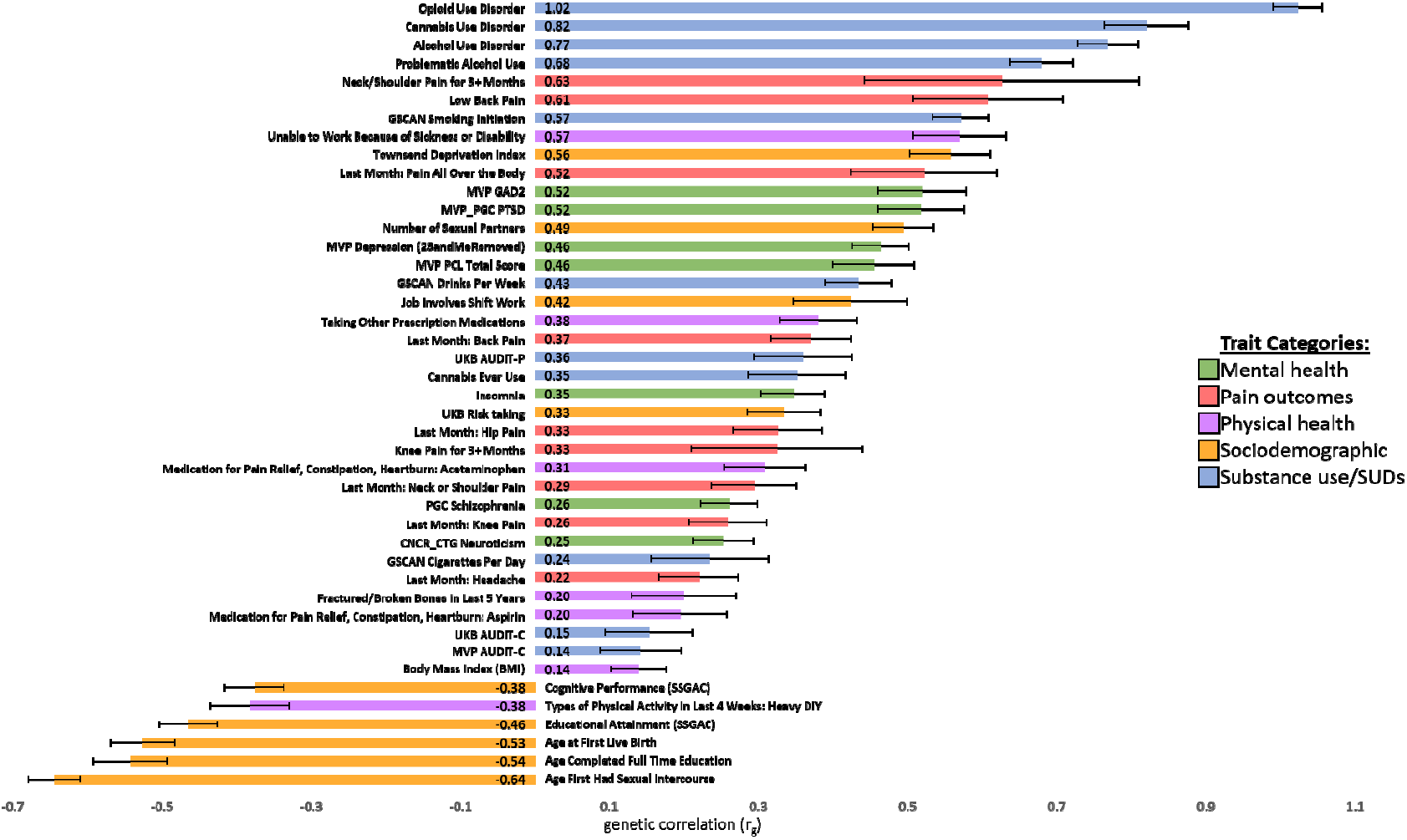
EUR OUD GWAS genetic correlations (r_g_) with mental health, pain, physical health, sociodemographic, and substance use traits of interest.

OUD also demonstrated statistically significant genetic correlations with many mental health, pain, physical health, and sociodemographic traits. The strongest positive correlations across the respective domains were with Generalized Anxiety Disorder (r_g_=0.52; *p*=2.89×10^−18^) and PTSD (r_g_=0.52; *p*=3.87×10^−19^), lower back pain (r_g_=0.61; *p*=1.22×10^−09^), inability to work due to being sick or disabled (r_g_=0.57; *p*=1.31×10^−20^), and scores on the Townsend Deprivation Index (r_g_=0.56; *p*=1.13×10^−25^). OUD was negatively genetically correlated with measures of sexual reproductive behavior (age of first sexual intercourse [r_g_=-0.64; *p*=4.43×10^−76^]), indices of educational attainment (age of school completion [r_g_= -0.54; *p*=9.41×10^−28^]) and cognitive performance (r_g_= -0.38; *p*=1.54×10^−20^), and levels of past month “Heavy Do It Yourself” physical activity (r_g_= -0.38; *p*=7.37×10^−13^), amongst others (**Supplemental Tables)**.

### Multi-trait analysis of European GWAS summary statistics (MTAG)

A multi-trait analysis using MTAG was supported by strong genetic correlation for OUD with CanUD (r_g_=0.82; *p* =1.14×10^−47^) and AUD (r_g_=0.77; *p* = 6.36×10^−78^) in EUR individuals. The OUD-MTAG analysis resulted in an increase in effective sample size from the original EUR OUD GWAS N_effective_=56,991 (GWAS mean □^2^=1.18) to an equivalent sample size of N=128,748 (GWAS mean □^2^=1.40). The increase in equivalent sample size and detection power from MTAG resulted in the identification of 18 independent GWS risk loci (**Figure 2**), some previously associated at either the variant level, or that reside in genes associated with, psychiatric and substance use outcomes in previous GWAS. The *FURIN* risk locus identified in the EUR OUD GWAS was GWS in the OUD-MTAG but *OPRM1* was not.

The top OUD-MTAG association was with rs11229119 (*p*=7.03×10^−11^) on chromosome 11 mapping to both *TMX2* and *CTNND1*. The second strongest was with *NICN1* (rs77648866; *p*=1.82×10^−10^) on chromosome 3. Additional GWS associations included *FOXP2* (rs1989903; *p*=2.47×10^−10^), *PDE4B* (rs7519259; *p*=2.68×10^−10^), *SLC39A8* (rs13135092; *p*=3.60×10^−10^), *NCAM1* (rs1940701; *p*=9.63×10^−10^), *RABEPK* (rs864882; *p*=1.24×10^−09^), *PLCL2* (rs55855024; *p*=7.89×10^−09^), and *FTO* (rs7188250; *p*=3.63×10^−08^). One of the *FURIN* variants identified in the EUR OUD GWAS was also GWS in the OUD-MTAG (rs17514846; *p*=2.30×10^−08^). The top *OPRM1* association was with rs1799971 (*p*=1.39×10^−06^). Of the 18 GWS loci, three were GWS in the AUD GWAS and one was GWS in the CanUD GWAS **(Table 4)**.

**Table 4.** Genome-wide significant (*p*=5.00×10^−08^) GWAS associations in OUD MTAG analysis.

| Chr | Pos | MarkerName | Allele 1 | Allele 2 | Gene(s) | Z | P-value | p-value in AUD | p-value in CanUD |
| --- | --- | --- | --- | --- | --- | --- | --- | --- | --- |
| 11 | 57535966 | rs11229119 | T | C | *TMX2-CTNND1* | -6.5 | 7.03E-11 | $p = 2.60\text{e-}07$ | $p=6.68\text{e-}05$ |
| 3 | 49469449 | rs77648866 | A | G | *NICN1* | 6.33 | 1.82E-10 | $p = 5.90\text{e-}07$ | $p = 5.89\text{e-}03$ |
| 7 | 114137940 | rs1989903 | A | G | *FOXP2* | -6.2 | 2.47E-10 | $p = 1.23\text{e-}03$ | **$p = 3.52\text{e-}09$** |
| 1 | 66434743 | rs7519259 | A | G | *PDE4B* | 5.73 | 2.68E-10 | $p = 9.94\text{e-}08$ | $p = 1.36\text{e-}05$ |
| 4 | 103198082 | rs13135092 | A | G | *SLC39A8* | 6.51 | 3.60E-10 | **$p = 4.898\text{e-}18$** | $p = 0.88$ |
| 11 | 112869404 | rs1940701 | T | C | *NCAM1* | -6.1 | 9.63E-10 | $p = 4.91\text{e-}06$ | $p = 1.67\text{e-}03$ |
| 9 | 127968109 | rs864882 | T | C | *RABEPK* | -5.6 | 1.24E-09 | $p = 2.45\text{e-}05$ | $p = 0.064$ |
| 6 | 19076417 | rs9350100 | T | C | *RP11-254A17.1* | 5.62 | 1.76E-09 | $p = 3.37\text{e-}05$ | $p = 2.89\text{e-}03$ |
| 1 | 91208451 | rs2166171 | T | C | *BARHL2* | -5.88 | 4.80E-09 | $p = 1.18\text{e-}06$ | $p = 2.76\text{e-}05$ |
| 3 | 16850764 | rs55855024 | A | C | *PLCL2* | 5.73 | 7.89E-09 | $p = 1.50\text{e-}06$ | $p = 0.20$ |
| 7 | 75622281 | rs6467958 | T | C | *TMEM120A* | -5.5 | 1.06E-08 | $p = 2.24\text{e-}05$ | $p = 4.02\text{e-}03$ |
| 11 | 113477081 | rs11214677 | T | C | *TMPRSS5* | -5.65 | 1.16E-08 | $p = 1.23\text{e-}04$ | $p = 1.65\text{e-}05$ |
| 1 | 28989020 | rs6667501 | A | G | *GMEB1* | 5.55 | 1.74E-08 | $p = 3.66\text{e-}04$ | $p = 2.21\text{e-}03$ |
| 19 | 45453763 | rs10422888 | A | G | *CTB-129P6.11* | 5.55 | 1.99E-08 | $p = 5.83\text{e-}05$ | $p = 6.22\text{e-}03$ |
| 15 | 91416550 | rs17514846 | A | C | *FURIN* | -5.59 | 2.30E-08 | $p = 2.85\text{e-}03$ | $p = 0.092$ |
| 15 | 47645174 | rs73403005 | A | G | *SEMA6D* | -5.69 | 2.46E-08 | **$p = 6.06\text{e-}09$** | $p = 2.38\text{e-}03$ |
| 13 | 96932868 | rs2389631 | A | C | *HS6ST3* | -5.61 | 2.53E-08 | $p = 8.72\text{e-}05$ | $p = 0.055$ |
| 16 | 53834607 | rs7188250 | T | C | *FTO* | 5.64 | 3.63E-08 | **$p = 4.41\text{e-}12$** | $p = 0.61$ |

The OUD-MTAG summary data was highly genetically correlated with the largest previously published GWAS of OUD to date at r_g_=0.98 (*p*=1.22×10^−77^)(7). All estimates of genetic correlation for the OUD-MTAG analysis can be found in **Supplemental Tables**.

### Phenome-wide Association Study (PheWAS)

The top PheWAS association for OUD was with substance addiction and disorders (OR=1.53; *p*=2.12×10^−69^). Additional top OUD associations included Tobacco use disorder (OR=1.26; *p*=3.38×10^−56^), chronic pain (OR=1.25; *p*=2.32×10^−28^), alcohol-related disorders (OR=1.35; *p*=1.04×10^−23^), mood (OR=1.13; *p*=1.27×10^−22^) and anxiety (OR=1.14; *p*=1.00×10^−21^) disorders, viral hepatitis C (OR=1.33; *p*=3.04×10^−20^), and suicidal ideation or attempt (OR = 1.49; *p*=2.17×10^−19^), amongst others **(Figure 4)**.

**Figure 4.**
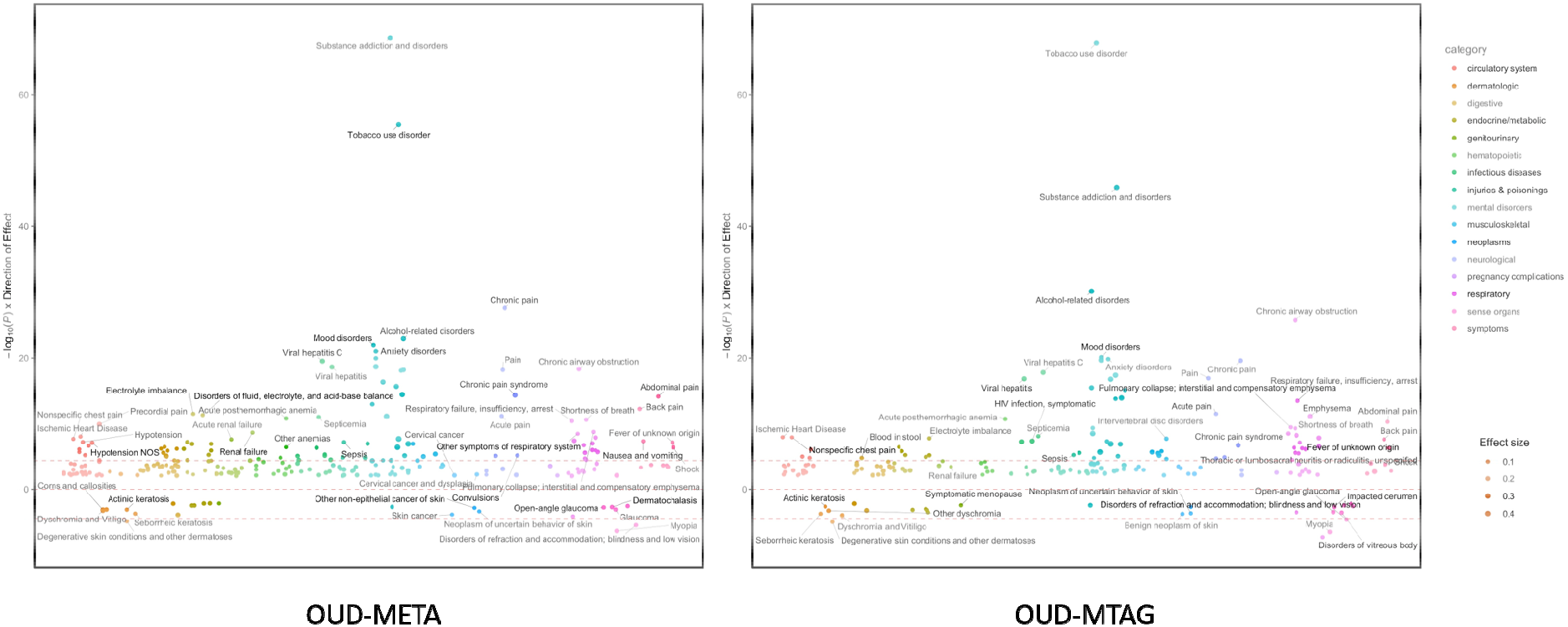
BioVU PheWAS results for EUR OUD GWAS (OUD-META; left panel) and OUD multi-trait analysis (OUD-MTAG; right panel) *Note*. Phenome-wide association study (PheWAS) results for 1291 clinical outcomes in BioVU. Y-axis represents the -log_10_(*p-*value) multiplied by the direction of effect. Diameter of data point corresponds to magnitude of effect size (i.e., larger dot=larger effect size). Data points below and above lower and upper red dashed line, respectively, indicate significant PheWAS association exceeding Bonferroni correction (*p*=3.87×10^−05^).

Very similar patterns of association were found for the OUD-MTAG PheWAS. The top associations were with Tobacco use disorder (OR=1.30; *p*=1.37×10^−68^), substance addiction and disorders (OR=1.42; *p*=1.15×10^−46^), and alcohol-related disorders (OR=1.42; *p*=7.36×10^−31^). OUD-MTAG also demonstrated significant associations with mood (OR=1.12; *p*=7.76×10^−21^) and anxiety (OR=1.13; *p*=1.45×10^−20^) disorders, chronic pain (OR=1.20; *p*=2.51×10^−20^), viral hepatitis C (OR=1.32; *p*=1.73×10^−18^), and suicidal ideation or attempt (OR=1.47; *p*=4.52×10^−18^) **(Figure 4)**. (Full description of PheWAS results: **Supplementary Tables)**.

## Discussion

We present a large genetic study of OUD, with an overall sample size of 639,709 (EUR=554,832; AFR=84,877) individuals (N_cases_=20,858 [EUR=15,423; AFR=5,435]). This study is the first to provide evidence of a GWS single-variant GWAS association between *FURIN* and OUD. We also support findings from previous OUD GWAS implicating *OPRM1* as a GWS risk factor for OUD(7), including the well-established *OPRM1* coding variant (rs1799971) and additional *OPRM1* associations that remained statistically significant in a cross-ancestry analysis of EUR and AFR populations. We add evidence of gene and gene-set associations with OUD, and provide robust estimates of OUD SNP-heritability and genetic correlations with many etiologically-relevant traits. Further, we apply a multi-trait approach for OUD genetic discovery utilizing the high degree of genetic correlation across SUDs (OUD, AUD, CanUD) to increase power, yielding an equivalent sample size of 128,748 and 18 GWS OUD-MTAG risk loci. PheWAS of OUD and OUD-MTAG demonstrated similar patterns of clinical associations across the phenome, suggesting that these traits are capturing similar phenomenon.

Compared to other complex psychiatric traits, there are comparatively small samples available for genetic analysis of individuals with drug use disorders, particularly those involving illegal substances (heroin, cocaine)(8,9). Thus, a strategy that increases statistical power by incorporating other sets of samples—for example, from GWAS of closely-related but non-identical traits such as other SUDs—could help advance our understanding of the genetic architecture of OUD. This study brought much more information to bear on the analysis of OUD risk variation, resulting in the identification of many more loci. These associations included three specific to OUD (*OPRM1* and *FURIN*) based upon findings from the EUR OUD GWAS, and 18 loci identified in the OUD-MTAG analysis (also including *FURIN*). The OUD-MTAG loci did not include any *OPRM1* variants identified in the OUD-specific analysis. This is surprising given the assumption that MTAG should specifically isolate OUD-related variance. The absence from the MTAG analysis any association mapped to *OPRM1*, a locus that should be highly-specific to OUD, is unexpected.

*FURIN* was associated with OUD risk in both SNP-based and gene-based analyses. *FURIN* (*Furin, Paired Basic Amino Acid Cleaving Enzyme*) encodes the endoprotease furin enzyme that serves a primary role in regulating synaptic neuronal activity, including the synthesis of brain-derived neurotropic factor and regulation of neurotrophin levels in the brain(26). Variation in *FURIN* has been associated with multiple psychiatric outcomes including schizophrenia(27,28) and studies examining genetic and phenotypic overlap between schizophrenia and bipolar disorder(29,30). The two top *FURIN* SNPs associated here with OUD are in strong LD (r^2^=0.91). The second strongest *FURIN* association in the current study, rs17514846, has been significantly associated with multiple cardiovascular and hypertension outcomes (31,32), and was also GWS in a GWAS of parents’ attained age (current age of parents or parental age at death)(33). A statistically-significant *FURIN* gene-level association being driven by rs17514846 was reported between *FURIN* and opioid addiction(34). In a targeted follow-up in the *FURIN* gene region, they also reported significant association between rs11372849 (lead SNP in the current study) and opioid addiction. Accumulating evidence linking *FURIN* and opioid outcomes, including the *FURIN* GWAS associations reported in the present study along with evidence of gene-based associations with opioid addiction(34) reflect the high degree of co-morbidity between OUD and psychiatric and physical health traits. Further investigation into *FURIN*’s involvement in the genetic underpinnings of physical health, psychiatric outcomes, and OUD is needed.

Our findings support previous OUD GWAS implicating *OPRM1* genetic variation as a risk factor for opioid addiction and OUD(7,34) and extend GWS findings for *OPRM1* as a risk factor in a cross-ancestry analysis of EUR and AFR populations. The top association in the EUR OUD GWAS was with the *OPRM1* coding variant (rs1799971), with an additional *OPRM1* variant (rs79704991) in low-LD with rs1799971 (r^2^=0.02) also identified. Two *OPRM1* variants were also found to be GWS in the cross-ancestry OUD GWAS (rs1799971 and rs9478500; r^2^=0.02). Rs9478500 was previously GWS for opioid addiction in EUR(34). There is well-documented evidence of *OPRM1*’s complex haplotype structure and the potential for multiple independent *OPRM1* risk loci influencing risk for OUD and SUDs(35). The conditional analysis of the top EUR OUD GWAS *OPRM1* variants (rs1799971 and rs79704991; r^2^=0.02) demonstrated that these variants are not independent. Future studies of larger cohorts with diverse ancestral backgrounds will be important for disentangling the effects of OUD risk loci across the *OPRM1* region, including those of the known-functional rs1799971 variant which may or may not be the variant motivating previous findings.

Estimates of heritability and genetic correlation also provide insight into the genetic etiology of OUD. We found a h^2^_SNP_ =12.75% (Z=11.28); a larger and more statistically robust estimate to the estimate reported in Zhou et al., 2020 (h^2^_SNP_ =0.113; Z=6.27), as would be expected from a ∼ 46% increase in EUR OUD cases. However, the comparison between these two studies is not direct: the largest previous GWAS(7) used opioid-exposed controls, while we used a broader control definition, including not only individuals who were opioid-exposed, but subjects with no OUD assessment. This was necessitated by the fact that many of the available datasets did not define exposed controls and would have been excluded had we used the exposed control definition. Findings from the current study do not establish whether the control definition impacted the detection of genetic loci or the genetic architecture of OUD.

OUD was positively genetically correlated with other SUDs (e.g., CanUD, AUD) and psychiatric conditions (e.g., PTSD, depression, schizophrenia), with lower correlations for measures of substance use (as opposed to dependence; e.g., AUDIT-C), suggesting that OUD is more akin to measures of substance dependence than use *per se*. OUD was genetically related to multiple forms of chronic pain (e.g., lower back pain) and indicators of impairment (inability to work, decreased physical activity) and significantly genetically correlated with socioeconomic hardship (Townsend Deprivation Index) and lower levels of educational attainment. These patterns of genetic correlation are consistent with high rates of co-occurrence of OUD with SUDs and psychiatric disorders in epidemiologic studies(36,37). Beyond epidemiologic estimates, SUDs and psychiatric disorders have also been demonstrated to be risk markers for severe opioid-related outcomes, such as opioid overdose(38). Educational attainment and economic hardship have also been associated with higher rates of opioid overdose and opioid overdose-related deaths(39). These patterns of genetic correlation are consistent with the complex clinical presentation of OUD and underscore the need for prevention and intervention in underserved populations and individuals with chronic and severe OUD.

We examined the utility of using MTAG to increase the information available from the limited number of genotyped OUD samples currently available. The OUD multi-trait analysis was feasible given the high genetic correlations with CanUD (r_g_=0.82) and AUD (r_g_=0.77) and increased by an order-of-magnitude the number of GWS risk loci detected. While this provides proof-of-concept for this approach given that many of the loci identified via OUD-MTAG have previously been implicated with psychiatric and substance use outcomes, *OPRM1* was not GWS in the OUD-MTAG analysis, so increased detection may have come at the cost of specificity for OUD. However, only 4 of the 18 OUD-MTAG GWS associations were GWS in the respective AUD and CanUD GWAS used as MTAG instruments, so the MTAG results did not simply reflect the findings from AUD and CanUD GWAS.

A PheWAS across 1,291 clinical outcomes also demonstrated convergent patterns of association between OUD and OUD-MTAG with common comorbidities (e.g., SUDs, psychiatric traits, chronic pain, viral hepatitis C), supporting that these two analyses capture genetic factors that underlie similar clinical presentations and related impairment. Additionally, summary data from the OUD MTAG analysis was highly genetically correlated (r_g_=0.98) with OUD(7), so it appears that the OUD-MTAG did capture genetic information relevant to OUD risk, though measuring the risk for OUD through a genetic liability for SUDs more broadly. That is, genetic risk for OUD may be a combination of a broader addiction liability (OUD-MTAG risk loci) combined with the opioid-specific genetic effects (e.g., *OPRM1*) that were found in the OUD single-trait analysis that are also influencing risk.

The distinction between substance-specific genetic effects and general SUD liability has long been of interest in genetic studies of SUDs. Quantitative genetic studies have demonstrated both substance-specific influences, as well as heritable factors that contribute to SUDs more broadly(40,41). Up to 38% of variation in opioid dependence was reportedly accounted for by opioid-specific factors that were not shared with other SUDs(42). Molecular genetic studies have begun to disentangle common vs. substance-specific genetic influences, reporting evidence to suggest the presence of a common unitary addiction factor that can account for risk across SUDs, in addition to substance-specific influences(43). Larger-scale studies of OUD will be needed to advance OUD genetic discovery and parse genomic influences specific to OUD from those underlying risk for SUDs more broadly. However, this will require many more genotyped OUD cases, because it cannot be accomplished via statistical methods alone.

The present study has limitations. Despite including all genotyped OUD subjects available to date, the OUD-only component of the present study is smaller than GWAS for other substance use behaviors (alcohol and nicotine use)(14) because OUD cases are underrepresented in available datasets. The MTAG analysis yielded a much larger sample, but at the apparent cost of a reduction in specificity marked by the non-significance of the *OPRM1* locus in the OUD-MTAG. To maximize sample size while maintaining OUD diagnosis to define case status in extant datasets, we used an unscreened control group, which although not optimal, allowed for the inclusion of additional cohorts that included OUD cases. Previous studies have provided evidence for important consideration being given to OUD control definitions(6,34). Additionally, an inadequate number of OUD cases and controls limited our ability to identify GWS variants contributing to risk of the disorder in population groups other than EUR. This must be addressed by purposeful recruitment of AFR and other non-EUR OUD subjects.

In conclusion, we report novel findings from a large-scale GWAS meta-analysis of OUD and explore multi-trait approaches to advance discovery for understudied traits such as OUD. These identified genomic risk factors for the development of OUD and underlying biology, and highlight the need to assemble large OUD datasets that include individuals from diverse ancestral backgrounds. To advance our scientific understanding of OUD risk will require study of a range of human traits (e.g., clinically diagnosed OUD and prescription painkiller use)(44). This will be necessary to improve further our understanding of the genetic etiology of OUD and translation of genetic findings to help address the opioid public health crisis and reduce preventable deaths.

## Data Availability

All data produced in the present study will be available upon request to the authors

