## Supplemental Methods for "Genome-wide association study and multi-trait analysis of opioid use disorder identifies novel associations in 639,709 individuals of European and African ancestry"

*Phenotyping*

All cohorts used in the present analysis were assessed for a lifetime diagnosis of OUD case status. OUD case status was determined via electronic health records (EHRs) or semi-structured interviews that assessed DSM-IV(1) OUD criteria **(Table 1).** Methods to define control status varied **(Table 1).** Controls were predominately unscreened individuals with no known OUD diagnosis; from the PGC-SUD GWAS(2) we selected the comparison with the opioid-unexposed control group.

*Previously unpublished GWAS cohorts*

Previously unpublished genome-wide association studies (GWAS) of opioid use disorder (OUD) from BioVU (3), iPSYCH (4), and Yale-Penn(5,6) (Yale-Penn 3; results from Yale-Penn 1 and 2 were published previously) were included in the present OUD meta-analysis. Study descriptions are provided below.

**BioVU OUD GWAS**

 We used de-identified clinical data from Vanderbilt University Medical Center’s biobank, BioVU. We included 4,665 individuals (933 OUD cases and 3,732 controls). Controls were matched on a 1:4 ratio based on sex, race, ethnicity, median age of the longitudinal EHR measurements, and density of records (number of ICD codes and length of record). OUD status was defined as having at least one OUD ICD code. OUD controls were defined as anyone without an OUD ICD code. Details on the quality control process have been described elsewhere(7).

Genotype data were generated using the Illumina Multi-Ethnic Genotype Array (MEGAEX) for 94,474 individuals. Genotypes were filtered for SNP(<0.95) and individual (<0.98) call rates, sex discrepancies, and excessive heterozygosity(|Fhet|>0.2)(8). The sample was then filtered for cryptic relatedness by removing one individual of each pair for which pihat>0.2. PCA using FlashPCA2 combined with CEU, YRI and CHB reference sets from 1000 Genomes Project Phase 3(9) was conducted to determine European Ancestry. We confirmed the absence of genotyping batch effects using ‘batch’ as the phenotype. We used the Michigan Imputation Server with the reference panel from the Haplotype Reference Consortium. SNPs were filtered for imputation quality (R2 > 0.3 or INFO > 0.95) and converted to hard calls. We restricted to autosomal SNPs with minor allele frequency >0.01. We removed SNPs that differed by >10% from the 1000 Genomes Project phase 3 CEU set(9) and those with a Hardy Weinberg Equilibrium *p*<10^-10^. The resulting data set contained hard-called SNP information for 9,386,383 SNPs in 72,828 individuals of European Ancestry. GWAS analyses were performed using SAIGE version 0.42.1(10) and including the top 10 genetic PCs as fixed effect covariates. The project was approved by the VUMC Institutional Review Board (IRB #160302, #172020, #190418).

**iPSYCH OUD GWAS**

**Case and control definition.** Those with any of the ICD diagnoses for opioid related disorders (F11.1 – F11.9) registered in the Danish hospital registers were considered as cases. The registers were followed up until Dec 2016. The controls were individuals from the iPSYCH cohort without any of the opioid related ICD diagnoses and aged at least 25 years by the end of register follow up. Individuals with and without psychiatric disorders were included in the controls. Comorbidity with psychiatric diagnoses was accounted for by including psychiatric diagnosis status for each of the disorders as binary covariates in the GWAS.

**Exclusion of related and non-European individuals.** After identifying individuals in the register with and without opioid related disorders, we excluded those who were not successfully genotyped, who were related and who were non-Europeans. The number of individuals removed at each step is shown in **Figure 1**. Analytical details on the removal of related individuals and non-Europeans can be found in Pedersen et al.(4).

**Down-sampling of controls.** Since including everyone in the iPSYCH cohort without opioid related disorders as controls would result in and extreme case control ratio, we down sampled the controls to have approximately 10 times the number of cases. In total, we included 379 cases and 5221 controls in the GWAS. The down-sampling was performed separately for each of the comorbidity groups in order to maintain a similar proportion of individuals with other psychiatric disorders between cases and controls. The sample sizes split by comorbid diagnoses are shown in **Table 1**.

**GWAS.** GWAS was performed using Plink using logistic regression analysis. We analyzed only variants that were retained after QC filtering (INFO>0.80, MAF>0.01, sample and variant missing rate <1% etc.) in both the iPSYCH-2012 and iPSYCH-2015i samples. This reduced the final number of variants, which was around 5 million. We used hard call genotypes for the GWAS analysis. Both the iPSYCH-2012 and iPSYCH-2015 genotype datasets were merged into a single genotype dataset and was used for GWAS analysis. This is our usual practice for GWAS where case numbers are low. In such scenarios, running GWAS separately in iPSYCH 2012 and iPSYCH 2015i is tricky due to very sparse covariates matrix that leads to too many NAs in the logistic regression. The covariates included age, sex, psychiatric diagnoses and first 10 PCs.

**Yale-Penn 3 OUD GWAS**

The Yale-Penn sample includes 11,332 genotyped and phenotyped individuals recruited across three phases (Yale-Penn 1, Yale-Penn 2, and Yale-Penn 3) according to time of recruitment and genotyping array used. For this study, data for Yale-Penn 1 and 2 was included via the PGC-SUD summary statistics. All cohorts were ascertained via recruitment at substance use treatment centers or targeted advertisements for genetic studies of cocaine, opioid, and alcohol dependence, resulting in a sample highly enriched for problematic substance use, as well as control subjects and relatives. All participants were assessed using the Semi-Structured Assessment for Drug Dependence and Alcoholism (SSADDA)(11,12). Previous analyses of OUD including Yale-Penn 1 and 2 have been published previously in studies (5,6), including those used in the PGC-SUD discovery sample (2) included in the present study; published data on Yale-Penn 3 was previously limited to use in part for replication analysis or as a target sample for polygenic risk score analyses(e.g., 6).

Yale-Penn 3 includes 3,026 genotyped and phenotyped Americans of European (EUR; N=1,986) and African (AFR; N=1,040) ancestry passing standard quality control. Genotyping was performed at the Gelernter lab at Yale University using the Illumina Multi-ethnic Global Array containing 1,779,819 markers, followed by genotype imputation using Minimac3 (13) and the Haplotype Reference Consortium reference panel (14) as implemented on the Michigan imputation server (https://imputationserver.sph.umich.edu).

For the present analysis, Yale-Penn 3 EUR (N_EUR_=1,986) and AFR (N_AFR_=1,040) participants were included. DSM-IV opioid abuse and dependence diagnoses based on SSADDA assessments were used to determine OUD case status (15,16). OUD controls were defined as anyone not meeting criteria for opioid abuse or dependence. Of the 1,986 EUR participants 22.56% met criteria for OUD (N_CASE_=448); 16.54% of AFR participants (N_CASE_=172) met criteria for OUD diagnosis.

*Gene and gene-set analysis*

GWAS summary data from the respective meta-analyses were used to carry out gene-based and gene-set analyses using MAGMA (Multi-marker Analysis of GenoMic Annotation)(17) as implemented in the FUMA platform (Functional Mapping and Annotation)(18). Single nucleotide polymorphisms (SNPs) were mapped to 16,113 protein-coding genes based upon physical position. GWS for the gene-based tests was defined via Bonferroni correction as *p*=3.10x10^-06^ (0.05/16,113). An analysis of gene-sets curated based upon gene function and biological pathways (MsigDB; *N=*15,458) was also performed, with GWS defined as *p*=3.23x10^-06^ (0.05/15,458).

**Table 1. IPSYCH OUD GWAS sample sizes split by comorbid diagnoses.**

|  | **With OUD** | **Without OUD** | **Down sampled controls** | **N in full final sample** |
| --- | --- | --- | --- | --- |
| Controls | 40 | 15580 | 400 | 400 |
| ADHD | 121 | 6131 | 1210 | 1468 |
| ASD | 17 | 3515 | 170 | 381 |
| Schizophrenia | 127 | 4023 | 1270 | 1555 |
| Bipolar | 34 | 2140 | 340 | 579 |
| MDD | 183 | 20429 | 1830 | 2889 |
| Anorexia | 15 | 2915 | 150 | 268 |

**Figure 1. IPSYCH OUD GWAS EUR sample inclusion pipeline**

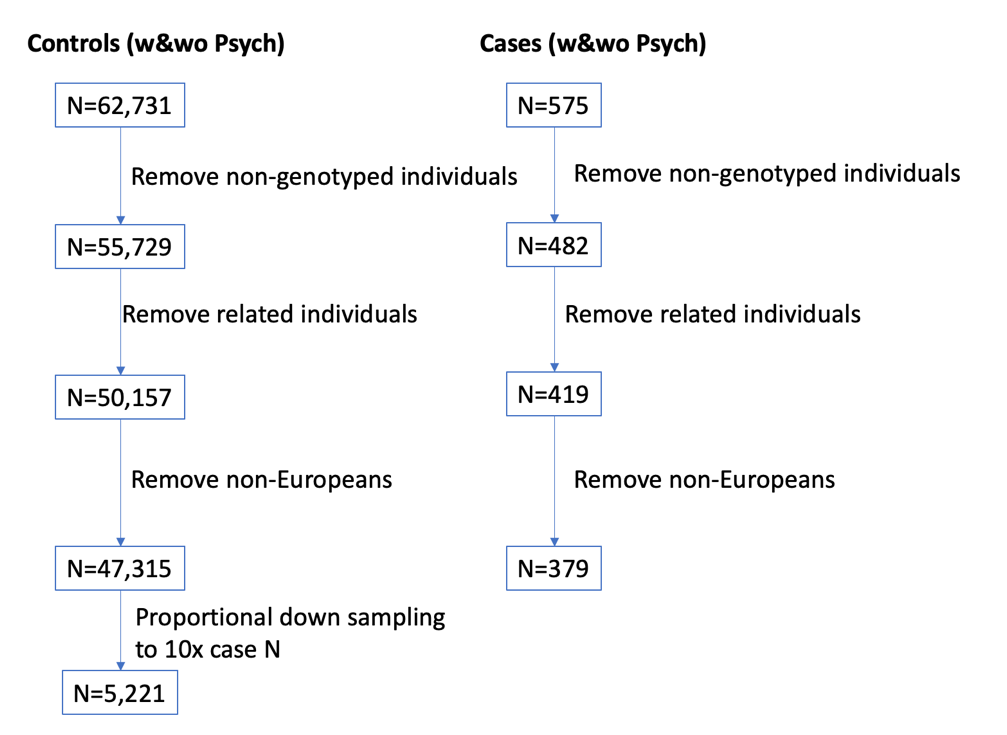
