## Supplemental Figures for "Genome-wide association study and multi-trait analysis of opioid use disorder identifies novel associations in 639,709 individuals of European and African ancestry"

**Supplemental Figure S1**. Regional plots of *OPRM1* gene region on chromosome 6 for EUR OUD GWAS top *OPRM1* associations (a) rs1799971 (*p*=4.92x10^-09^ ) and (b) rs79704991 (*p*=1.11x10^-08^; r^2^=0.02).

(a)

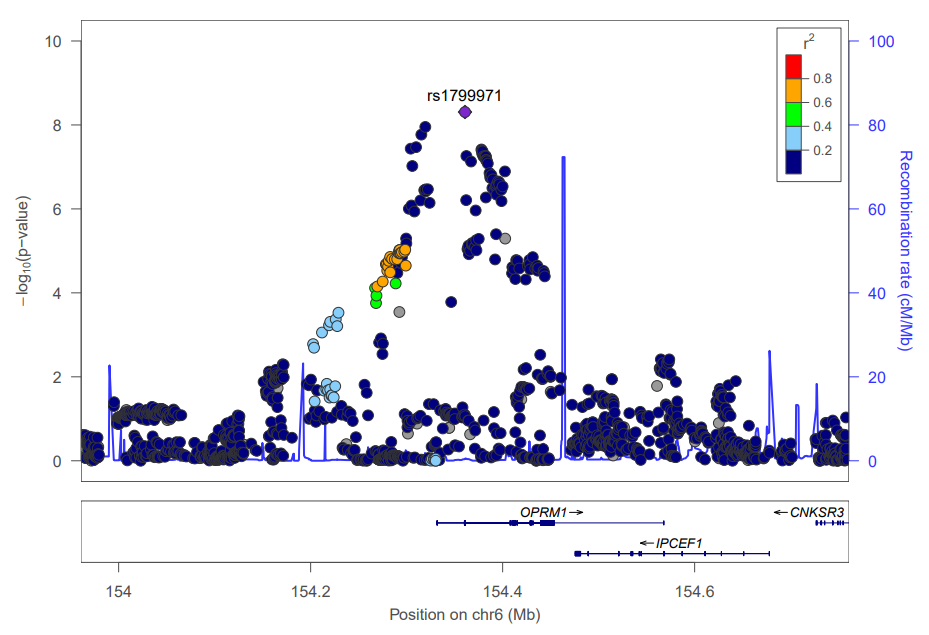

(b)

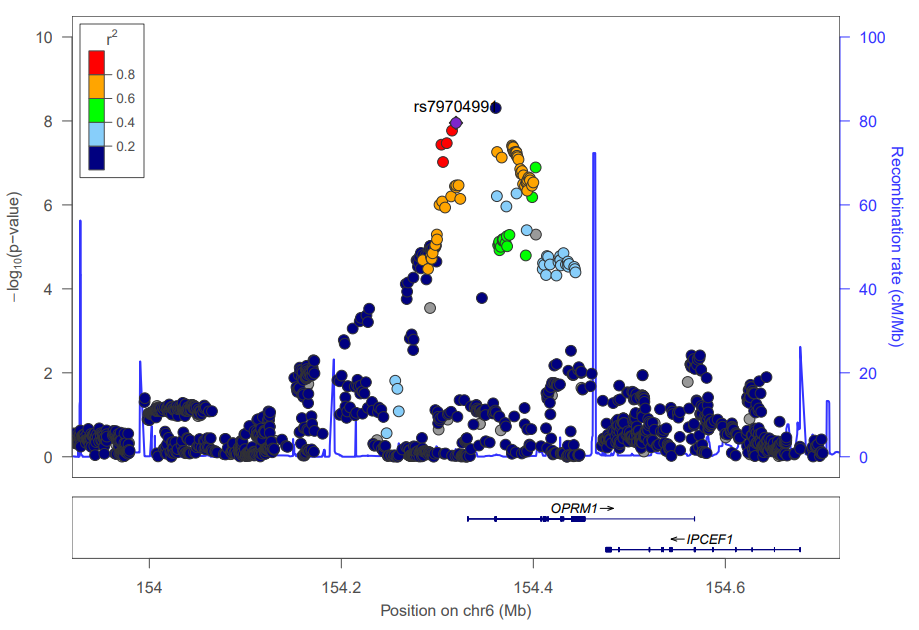

**Supplemental Figure S2.** Manhattan plot of African-ancestry OUD GWAS results

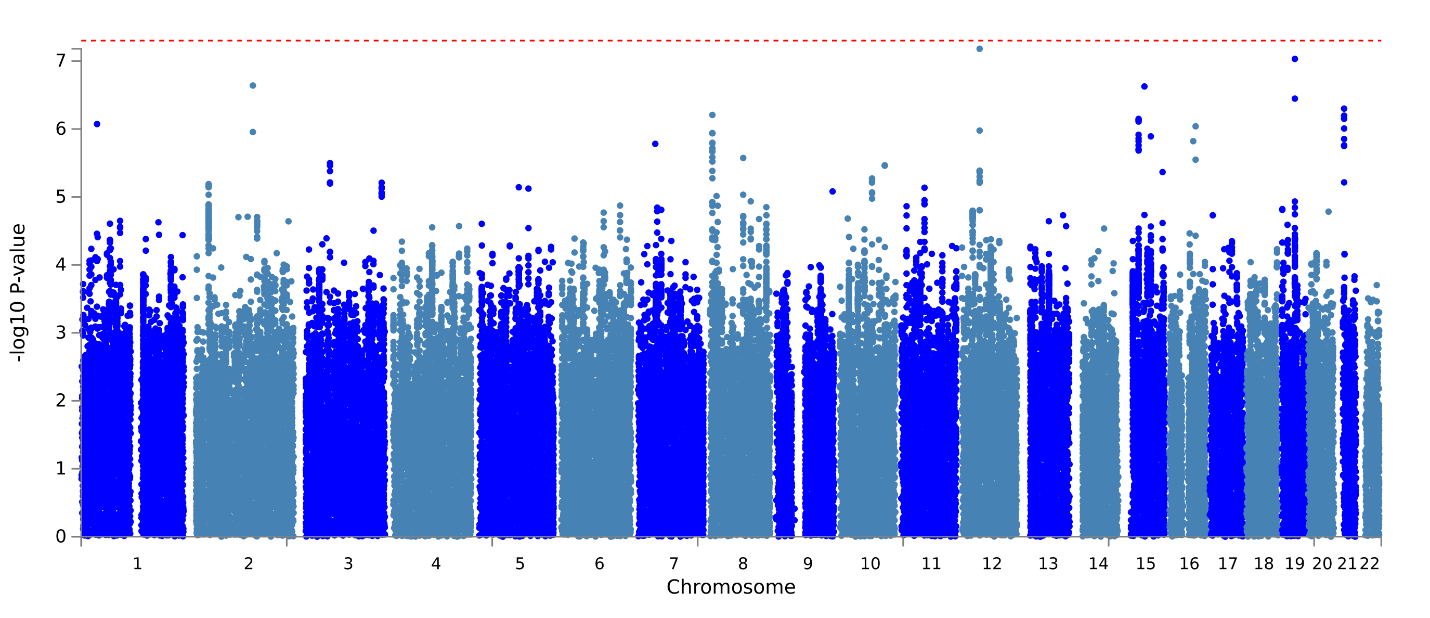

**Supplemental Figure S3.** Manhattan plot of AFR/EUR cross-ancestry OUD GWAS results.

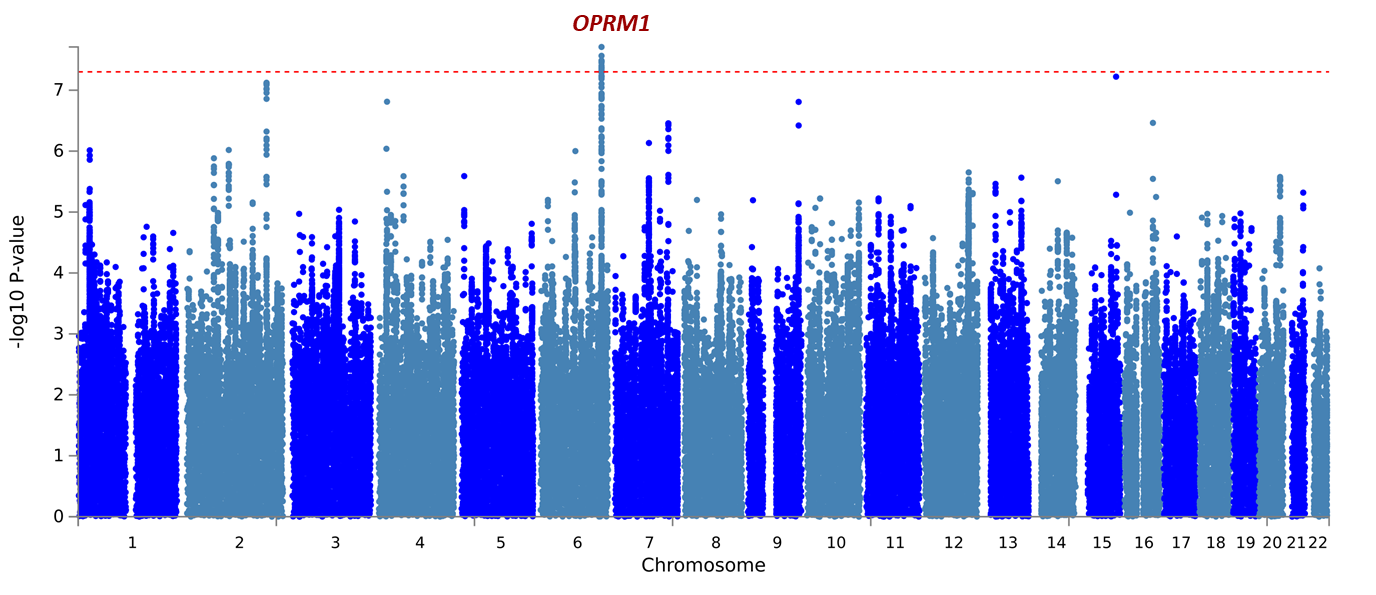

**Supplemental Figure S4.** Manhattan plot of EUR ancestry OUD gene-based results.

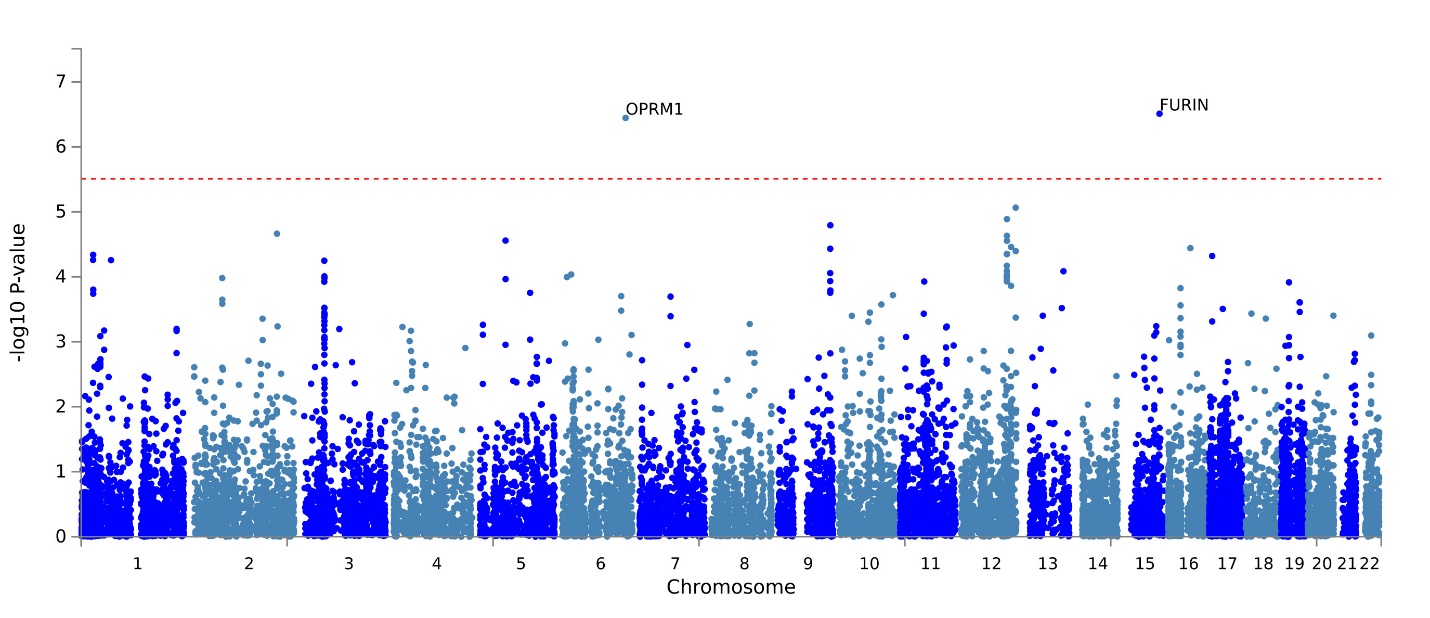

**Supplemental Figure S5**. Manhattan plot of AFR/EUR cross-ancestry OUD gene-based results.

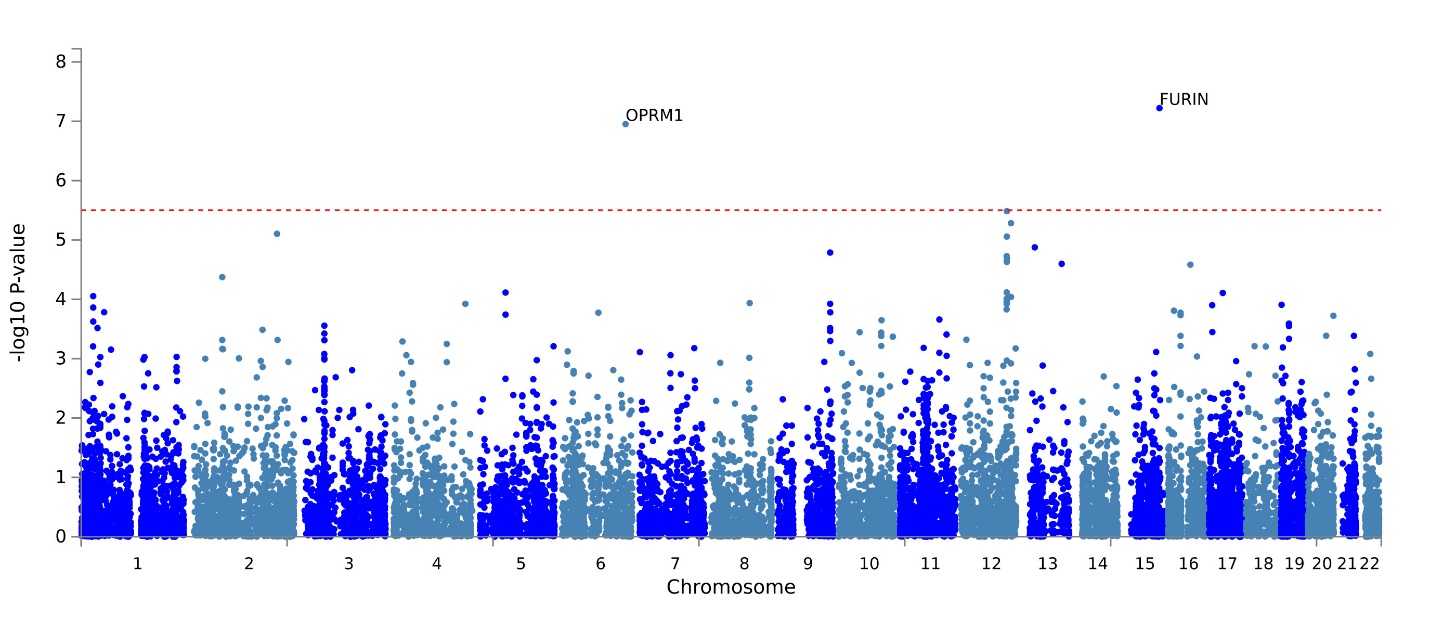
